# Psychosocial attributes of housing and their relationship with health among refugee and asylum-seeking populations in high-income countries: systematic review

**DOI:** 10.1101/2022.12.04.22283020

**Authors:** Tessa Brake, Verena Dudek, Odile Sauzet, Oliver Razum

## Abstract

**Introduction:** Housing as a social determinant of health should provide not only shelter, but also a feeling of home. We explored psychosocial pathways creating a sense of home and influencing the relationship between housing and health among asylum seekers and refugees (ASR) in high-income countries.

**Methods:** We performed a systematic review. To be included, studies had to be peer-reviewed, published between 1995 and 2022, and focus on housing and health of ASR in high-income countries. We conducted a narrative synthesis.

**Results:** 32 studies met the inclusion criteria. The psychosocial attributes influencing health most often identified were *control*, followed by *expressing status, satisfaction*, and *demand*. Most attributes overlap with material/physical attributes and have an impact on ASR’s mental health. They are closely interconnected with each other.

**Conclusions:** Psychosocial attributes of housing play an important role in the health of ASR; they are closely associated with material/physical attributes. Therefore, future research on housing and health of ASR should routinely examine psychosocial attributes, but always in association with physical attributes. The connections between these attributes are complex and need to be further explored.

**PROSPERO registration number:** CRD42021239495

## Introduction

The number of forcibly displaced people has been increasing in recent years. At the end of 2021, 89.3 million people were forcibly displaced. This number included 4.6 million asylum seekers and 27.1 million refugees [1]. In the coming years, further large refugee flows are expected due to extreme weather events as a result of climate change [1]. With these changes in climatic conditions and also demographic patterns, the social determinant of housing is becoming increasingly important for the health of the population [2]. Groups marginalized by, for example, socioeconomic status and/or race/ethnicity, not only have poorer health, but also experience poorer conditions with respect to social determinants, such as poorer access to adequate housing [3]. These groups, including ASR, are particularly affected by inadequate or insecure housing or outright denial of housing [2]. Poor housing is a major cause of health disparities, i.e., systematic differences in the health status of different population groups [2]. Addressing housing as a social determinant of health is relevant for improving health and reducing health disparities [4]. Adequate housing is a key indicator for successful integration [5].

### Housing as a social determinant of health

Many associations between housing and health can already be found in the empirical literature [2,6,7]. However, the literature also indicates that the causal relationships going beyond the direct effects of structural deficiencies of the dwelling are poorly understood [8-10].

According to the WHO definition [2], healthy housing should provide “*a feeling of home, including a sense of belonging, security and privacy*.” [2, p. 2]. A recent systematic review found significant associations with physical and mental health for a range of context-related attributes of housing among ASR, most of which were material/physical in nature. This relation was intertwined with other attributes, such as issues of discrimination, and the authors point to the need for better research tools to examine this link more generally [11].

The way these attributes are linked is often referred to as psychosocial pathways, which are considered important mediators in the relationship between housing and health [8-10,12]. We consider these attributes, which reflect on the social meanings associated with housing and create a sense of home, as psychosocial attributes of housing in our research. Figure 1 provides an overview of these relations. They serve as the starting point for our study. A challenge is to better define the concept of “home” and to make its relevant attributes measurable [13].

**Figure 1:**
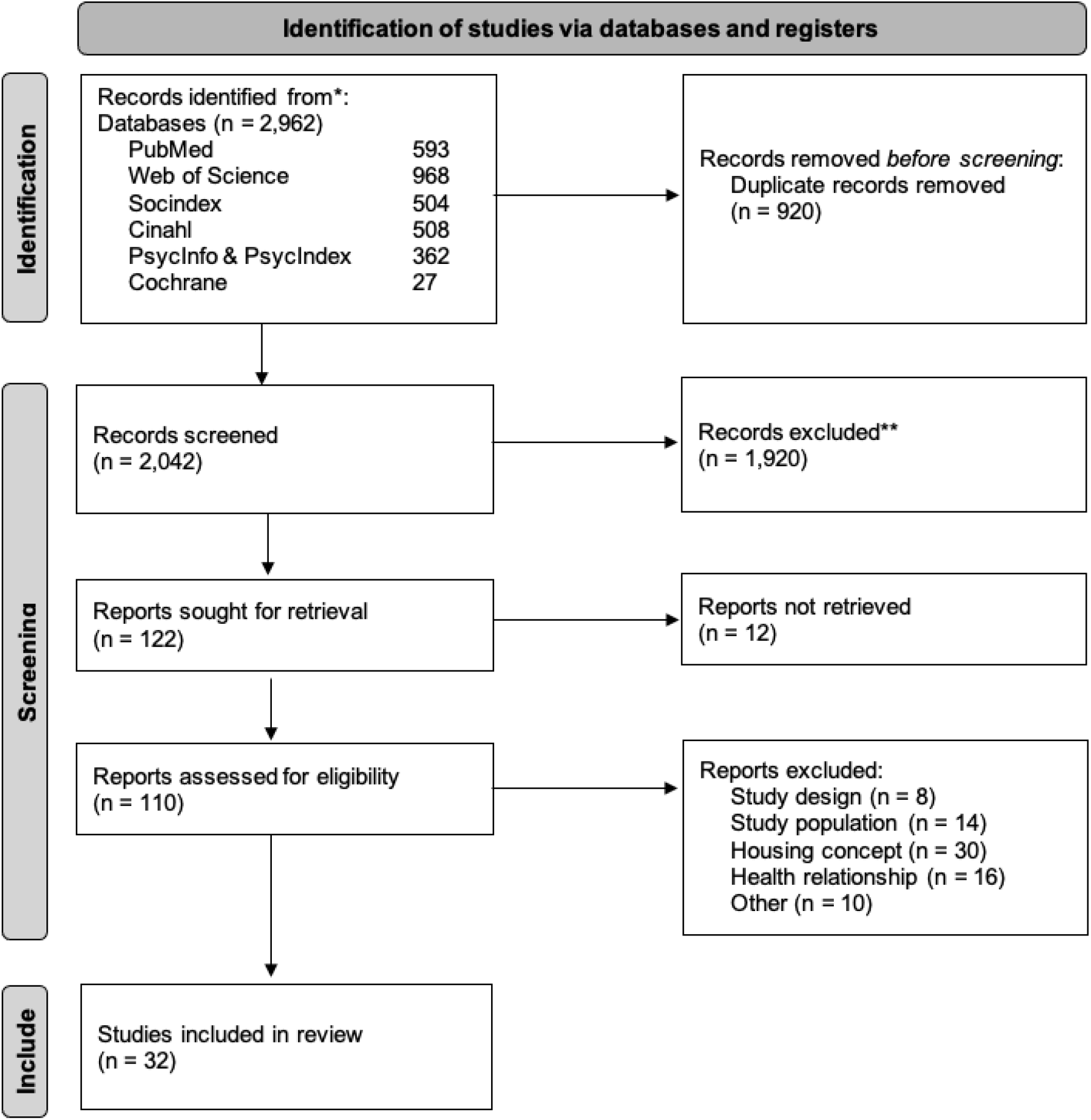
PRISMA Flow chart (Psychosocial attributes of housing and health among refugees, systematic review, high-income countries, 1995-2022)

### Psychosocial attributes of housing

To identify psychosocial attributes of housing, we refer to a framework proposed by Dunn [14]. Dunn collates such attributes in the “meaningful dimension of housing”, one of three key housing dimensions through which social inequalities and health consequences are produced. The dimension is conceptualized around three psychosocial attributes derived from the field of workplace organization: personal *control* (or decision latitude, the individual’s control over his/her daily tasks), *demand* (a measure of stress sources resulting from heavy workloads), and *social support* (and the expression of social status). Following the Job-Demand Control Model [15], individuals with more mentally demanding jobs and lower personal control are more likely to show mental distress. Social support (or increased isolation) can increase the impact of demand and control on health [16]. Considering this, Dunn [14] assumes that these attributes that are most important for health at work will also apply to the place where people spend most of their time: the home. Dunn adds a specific attribute: the satisfaction with living conditions. The psychosocial attributes are again each defined by indicators (see supplementary file 1).

Dunn did not focus on particular population subgroups in this work. We here aim to apply it to ASR and explore the psychosocial attributes of their housing and its role for health. We posit that these attributes present pathways helping to explain the association between material/physical attributes of housing identified in empirical research and health. To examine our postulate, we address the following research questions: *(1) To what extent are psychosocial attributes of housing taken into account in empirical research assessing the relationship between housing and health of ASR?* and *(2) What is the relationship between psychosocial attributes of housing and the health of ASR?*

## Methods

We conducted a systematic literature review, following the Preferred Reporting Items for Systematic Reviews and Meta-Analysis (PRISMA) checklist [17]. The protocol has been registered with PROSPERO (CRD42021239495).

### Searches

We applied the PECO (Population - Exposure - Comparison - Outcome) framework [18]. The population of interest included adult ASR who resettled to a high-income country. We set no limitation to gender, ethnicity, or time since migration. The exposure was related to the housing context and was assessed in two stages. First, we considered any material/physical attributes of housing assessed in the studies and their link to health to identify different categories of housing-related attributes. Second, we assessed which of these attributes falls under psychosocial attributes of housing according to Dunn [14]. In order not to exclude potentially relevant studies due to a too narrow concept of health, we included studies assessing any health-relevant outcome referring to physical and mental health, morbidity, and mortality, corresponding to the WHO definition of health [19]. We searched in PubMed, Web of Science, PsycINFO and PsycINDEX, SocINDEX and the Cochrane Library. We built the search terms (see supplementary file 2) by drawing on the PECO framework. We derived the specific terms from previous housing and/or forced migration research and conducted a pilot search in PubMed.

### Eligibility criteria

Inclusion and exclusion criteria are displayed in Table 1. We considered the year 1995 as a time point by which the consequences of the wars that ended in Bosnia and Croatia were increasingly thematized, as preliminary analyses in PubMed showed. We set no limitations regarding the language of the publication. Since attributes of housing are measured in different ways in empirical research, we kept eligibility criteria broad and included all studies that may point to any relationship between housing and health in the abstract or title by comprising the terms “housing” (or the synonyms: “dwelling”, “shelter” or “accommodation”) and “health” (or related: “morbidity”, “mortality”, “depression”, “anxiety” or “post-traumatic stress”). We excluded studies that focused on homelessness only or defined the housing context only broadly which would not provide insights into which housing-related attributes are potentially related to health. We did not include studies conducted on internally displaced persons or the general population of (im-)migrants unless the study provided disaggregated data for ASR.

**Table 1:** Eligibility criteria (Psychosocial attributes of housing and health among refugees, systematic review, high-income countries, 1995-2022)

| Inclusion criteria | Exclusion criteria |
| --- | --- |
| Peer-reviewed qualitative, quantitative or mixed-methods study published between 1995 and 2022 | Focus of the study is on homelessness only |
| Abstract or title includes the term housing or related synonyms (dwelling, shelter, accommodation) and health (or related terms morbidity, mortality, depression, anxiety, post-traumatic stress). | Housing concept is too broad or unspecified (i.e., if studies did not assess at least one subcomponent of housing) |
| Study must be conducted in HIC | Studies in which target group is comprised of internally displaced persons or general |
|  | population of (im-)migrants and data for ASR is not disaggregated |
| Target population must be ASR (forcibly displaced population) |  |

### Data selection and data collection process

Two reviewers (TB, VD) independently screened titles and abstracts of all search matches for eligibility. Next, we retrieved texts of potentially eligible studies and again assessed them independently. We discussed inconsistencies or disagreements, if necessary, with the help of a third reviewer from the study team.

For data extraction, we used a standardized, predesigned form which included aspects such as bibliographic information, study design and focus of the study, setting and sample characteristics, housing characteristics, health measures and main housing-related health outcomes. We piloted the form by having both reviewers extracting five (approx. 16%) of the studies. After discussing and clarifying inconsistencies in extraction, both reviewers extracted half of the remaining studies.

To identify psychosocial attributes of housing as objectively as possible, we used indicators derived from Dunn [14] (see supplementary file 1). We identified these in three ways: psychosocial attributes were directly assessed quantitatively, directly described by the study participants, or indirectly derived from the study results. The latter was the case when study participants referred to certain attributes without explicitly naming them, or when in quantitative research, variables measured aspects which were very similar to the indicators of attributes.

### Study quality assessment

We were aware that psychosocial attributes of housing are less tangible than material/physical attributes. We thus did not specify a certain concept of housing before the screening to avoid bias resulting from rejecting potentially relevant studies in the selection process. For the same reason, we included qualitative study designs since we assumed that psychosocial attributes of housing may be likely to emerge from perceptions and experiences of the target populations. We used the Mixed-Methods Appraisal Tool (MMAT) for critical appraisal of the study quality [20,21]. The MMAT provides different sets of criteria for assessing the methodological quality of different study designs: qualitative, quantitative, and mixed methods design. Two reviewers (TB, VD) independently applied the MMAT on half of the included studies after a pilot phase in which both assessed five studies (approx. 16%) and discussed inconsistencies.

### Data synthesis

We conducted a narrative synthesis since we expected a high heterogeneity of results and a diverse range of methods applied. We followed the Guidance on the Conduct of Narrative Synthesis for Systematic Reviews [22]. We conducted a thematic analysis to identify main housing-related attributes considered relevant for health. Using vote-counting, we investigated how many studies assessed material/physical and psychosocial attributes. To explore relationships within and between studies, we drew a conceptual map that depicts the relationships identified between housing-related attributes and health, also including findings of psychosocial attributes (see supplementary file 3). For assessing the robustness of the synthesis, we critically reflected on the synthesis process [22]. We discussed limitations of the data synthesis method, quality and generalizability of the evidence, and discrepancies identified in the findings and how we handled them.

## Results

We included 32 studies in the review (see supplementary file 4). Of these, 15 were of qualitative design and 17 of quantitative design. Of the quantitative studies, nine were of longitudinal, seven of cross-sectional design and one was a comparison between a cross-sectional and a longitudinal study. Figure 1 shows the PRISMA flowchart.

### Overview of studies

The largest proportion of studies was conducted in the UK (*n*=8) [25,27,28,39,47,48,50,52] followed by Germany (*n*=6) [24,31,34,42,45,51] and Australia (*n*=5) [29,30,32,36,54]. One study each was conducted in Canada [23], the Netherlands [26], Austria [37], Ireland [43], Switzerland [44], Norway [46], and Sweden [49]. Most studies (*n*=25) included adult ASR [23-25,27-29,31,32,34-36,39-41,43-45,47-54], and were published between 2015 and 2022 (*n*=24) [23-25,28,29,31,32,34-40,42-46,49-51,53,54]. The number of participants in qualitative studies ranged from 6 to 106, and in quantitative studies from 105 to 5,678 persons. 19 studies focused on the outcome of mental health [23,26-29,35-37,40,41,43,45-47,49-53]. We found housing as a main topic in seven studies [26,27,31,32,35,52,54], in the remaining 25 it was a side topic. The most common regions of origin of the participants were Syria, Afghanistan, Iraq, Iran, Somalia, and Pakistan.

After the risk of bias assessment, we classified 27 studies (84.4%) [23,24,26-29,31-37,39-45,47-52,54] as high and five (15.6%) [25,30,38,46,53] as moderate regarding their quality. None was of low quality. In the subset of quantitative-descriptive studies, the criterion “non-response bias” was not met in ten of 16 studies. “Representativeness” was not fully achieved in two studies. In one study, we awarded zero points for the sampling strategy. Three out of the 16 quantitative-descriptive studies were of moderate quality, the others of high quality. One quantitative non-randomized study was rated as of moderate quality because it did not meet the criteria of appropriate measures and complete outcome data. The evaluation of the qualitative studies showed different limitations. The criterion most frequently violated was the link to the research question. All qualitative studies were of high quality, except one of moderate quality.

### Relationship between housing and health among ASR from a psychosocial perspective

We identified seven categories of housing-related material/physical attributes: 1) crowding, space, and privacy, 2) housing instability (including: security of tenancy, affordability, and residential mobility), 3) safety, 4) physical conditions of/in the dwelling, 5) housing satisfaction, 6) neighborhood and location, and 7) institutional practices in the dwelling or accommodation center.

We found *crowding, space, and privacy* in 17 (53.1%) [24-16,31,32,34,37,39-42,44,48,50,51,53,54] and *housing instability* in 16 (50%) [27,29,30,34-37,40,41,47-50,52-54] of the 32 studies. We further identified the category *physical conditions* in nine (28.1%) [32-34,37-39,44,48,54] and *safety* in eight studies (25.0%) [31,32,34,39,40,48,50,54]. We identified fewer housing-related attributes in quantitative studies. Only in terms of *housing satisfaction* quantitative approaches dominated (23.5% quantitative vs. 6.7% qualitative; number referred to: 17 quantitative and 15 qualitative studies in total).

Psychosocial attributes were directly assessed quantitatively in 12.5% (*n*=4) [23,28,45,48] of the 32 studies, reported by the study participants in eight studies (25%) [32,34,39,41,43,50,52,54], and indirectly derived from the study results by the present reviewers in ten studies (31,3%) [25,31,35-37,40,44,46,48,50]. In general, we determined that psychosocial attributes were present in 20 (62.5%) [23,25,28,31,32,34-37,39-41,43-46,48,50,52,54] of the 32 studies.

Looking at the thematic analysis (see supplementary file 5), we found some overlap between the material/physical attributes and the psychosocial attributes according to Dunn [14]. We classified the material/physical attributes *lack of privacy and space* as attributes of the category *crowding, space, and privacy* under the psychosocial attribute of control. The material/physical housing-related category *housing instability* included attributes such as *residential mobility* and *housing costs/affordability*. As participants perceived these as burdens, we assigned *affordability* to the psychosocial attribute of demand, specifically to its indicator *strain of meeting costs*, and *residential mobility* to the psychosocial indicators *worry of frequent move* and *worry of forced move* as part of the psychosocial attribute of control. The category *safety –* within the shelter due to (aggressive) strangers and in the neighborhood – was often linked to a fear of crime and victimization. Thus, we assigned it to the corresponding psychosocial indicator *fear of crime/victimization*. Regarding neighborhood, good social relations were also associated with a sense of *belonging* as an indicator of the psychosocial attribute *expression status*. In one study, participants perceived the access to green spaces, as an aspect of *location*, as positive for health. We associated this, in turn, with the feeling of well-being at home and thus assigned it to the psychosocial indicator *place of refuge*. The fourth housing-related category *physical conditions of/in the dwelling* included attributes such as *gardens and outside spaces* which participants saw as a burden. We thus subsumed it under the indicator *housework strain*, as an indicator of the psychosocial attribute *demand*. In addition, *satisfaction* with housing emerged as a material/physical attribute, which is also defined as a psychosocial attribute according to Dunn (2002). The seventh housing-related category of material/physical attributes was *institutional practices in the dwelling*. When it came to low control regarding *institutional practices*, participants did not feel as comfortable as they would in a “real” home, which we, in turn, attributed to the psychosocial control indicator *place of refuge*. Moreover, participants felt ashamed by these practices and thus their home did not represent who they really are [43]. This was consistent with the definition of the psychosocial attribute of expressing status, specifically with its indicators *pride* and *self-reflection*.

The attributes we identified most frequently related to *control*. Of its four indicators, the indicator *place of refuge* was most prevalent (*n*=13, 40.6%) [25,31,32,34,39-41,43,44,46,50,52,54], followed by *fear of crime and/or victimization* (*n*=7, 21.9%) [31,32,34,35,39,40,54], and the indicators *concern about forced* [32,50,52,54] and *frequent moves* [36,37,40,52] (*n*=4, 12.5%). The attribute *expressing status* were the second most common. While we found its indicators *pride* [43] and *self-reflection* [43] once each (3.1%) in the 32 studies, we identified the sense of belonging to the neighborhood six times (18.8%) [31,32,35,46,48,54]. This attribute was followed by the attribute *demand*. Its indicator *strain of meeting costs* was addressed in 12.5% (*n*=5) [32,40,41,54], while the indicator *housework strain* was addressed in 3.1% (*n*=1) [32] of the studies. Finally, we identified the psychosocial indicator of *satisfaction* as part of the general attribute in five studies (15.6%) [23,28,31,45,48]. We identified all indicators except *satisfaction* more frequently in qualitative than in quantitative studies.

### Relationship between psychosocial attributes of housing and the health of ASR

#### Control

We identified the indicator *place of refuge* most commonly regarding health outcomes of ASR. A lack of control over the home [25,31,34,43,46,53,54] and/or the lack of a private retreat [25,32,34,39-41,44,50,54], including a safe place with enough space, negatively affected mental health. Having to share space with others and the resulting lack of privacy led to feelings of anxiety, discomfort, frustration, disempowerment, and sadness [34,40,41,53]. Participants perceived the lack of control in the sense of influencing daily decisions as a psychosocial stressor for both mental and physical health [43]. Other frequently encountered indicators of psychosocial attributes were *worry/strain of forced* and *frequent moves*. We found problems with forced moves in four studies, all of which related to a lack of control over the home, leading to disempowerment which subsequently affected mental health, or health and well-being in general [32,50,53,54]. We also identified four studies that pointed to issues related to frequent moves; in all of them, frequent moves were seen as stressors for health [36,37,40,53]. The last indicator referring to *control* pointed to *issues of victimization* and *fear of crime*. While four studies had identified fear of crime due to other residents in the accommodation facility [34,35,39,40], five referred to safety aspects in the neighborhood environment [31,32,35,39,54]. Fear affected mainly mental health, manifesting as anxiety, but also physical health through lack of sleep and stress [31,32,39,54]. Fear of violence, and witnessing neighborhood and interpersonal violence, were also important causes for involuntary moves [35].

#### Expressing status

The second most important attribute regarding the health of ASR was *expressing status*, especially the indicator *belonging*, which six studies addressed. Feelings of trust and confidence in the neighborhood contributed to feelings of belonging, which ASR perceived as positive for health and well-being [32]. The experience of discrimination in the neighborhood, in contrast, negatively affected health and at the same time strengthened the intention to move [35]. ASR who were less often in contact with locals had significantly poorer mental and physical health scores [31], as the feeling of social isolation led to depression and stress [48].

#### Satisfaction

Associations between satisfaction with the home and the health of ASR were reported in five studies [23,28,31,45,48]. According to a Canadian longitudinal study, low satisfaction with housing conditions was significantly associated with a severe and/or moderate level of depressive symptoms [23]. In addition, decreased satisfaction with accommodation was significantly associated with higher odds of poor emotional well-being [28]. Dudek et al. [31] found that ASR living in collective accommodation showed the lowest satisfaction with the living situation and thus had significantly poorer mental and physical health scores compared to ASR living in private accommodations.

#### Demand

The last material/physical attribute which we assigned to the housing related category *housing instability* and to the psychosocial demand indicator *strain of meeting costs* was *affordability*. We identified four studies which examined the effect of *affordability* on the health of ASR [32,40,41,54]. Participants considered the cost of living a fundamental burden causing stress and worry [54] and perceived it as a strain in terms of personal control, hindering them from moving from inappropriate to better housing [32].

## Discussion

We explored the psychosocial attributes of housing among ASR and their role for health, based on the assumption that the social meanings of home present pathways explaining the relationship between material/physical attributes of housing and health.

We identified seven housing-related categories of attributes relevant to health, ranging from *physical conditions, crowding, space, and privacy, safety, housing instability, neighborhood/location, institutional practices* up to the *satisfaction* with the home. We could assign most of the material/physical attributes to the categories *crowding/space/privacy* and *housing instability* which may indicate a high importance of these categories for ASR in HIC. The psychosocial attributes we identified most often referred to *control*, in particular the indicators *place of refuge* and *forced/frequent moves*. These indicators were also consistent with the two largest categories of material/physical attributes identified: *crowding, space and privacy*, and *housing instability*. From a health perspective, it seems to be important to have a private retreat where one feels comfortable and considers one’s own. These feelings of home are difficult to develop in overcrowded, instable accommodation. Due to the lack of control through institutional practices, the indicator *place of refuge* is also closely linked to the category *institutional practices in the dwelling*. Additionally, *fear of crime and violence* were major causes for anxiety, underpinning the housing-related category *safety* – in the accommodation facility, as well as in the neighborhood. We identified the material/physical attribute *affordability* as part of the category *housing instability* second most frequently. Participants of all studies perceived this attribute as a burden. This in turn reflects the indicator *strain of meeting costs*, which falls under the psychosocial attribute *demand*. We identified the strain of affordability as a fundamental problem that causes stress and worry. The attribute *expressing status* mainly referred to the indicator *belonging*, again underpinning the importance of neighborhood. But even within accommodations a sense of belonging can be fostered or impeded by *institutional practices*, with potential impacts on health. On the one hand, social support is available, but on the other hand, entry and exit controls, for example, can restrict the people living there. In contrast, the psychosocial indicators *housework strain, pride*, and *self-reflection* seemed to be less relevant. Lower or no *satisfaction*, as both a material/physical attribute and psychosocial attribute of housing, with current accommodation was significantly associated with a higher likelihood of poor emotional well-being with severe and/or moderate depressive symptoms.

### Limitations and strengths

Our study has limitations and strengths. Starting with limitations, we did not conduct a “grey” literature search and did not include other than peer-reviewed articles. This may have incurred publication bias [55] as non-significant results may not have been published [56]. If this were indeed the case, we would be overestimating the strength of association between psychosocial attributes and health of ASR. Because one of the three methods we used to identify psychosocial attributes was to derive them indirectly from study results, we may have been too subjective in our assessment. This could affect the reproducibility of our approach. Contrary to recommendations [20], the two reviewers only assessed five studies independently and discussed the results. The remaining studies were split up and assessed by one person each. This may have resulted in slightly different assessment of studies. We tried to minimize this problem by frequently communicating about our assessment procedures.

We included studies featuring a variety of methods and with heterogeneous results in our review. So, we could conduct only a narrative synthesis, including thematic analysis, instead of a meta-analysis, which may have reduced transparency and methodological clarity [57,58]. The heterogeneity of studies in terms of design, sample size, methodical approach, and research focus limits their comparability, interpretation, and generalizability.

However, our study also has notable strengths. To our knowledge, our review is the first to consider the psychosocial perspective as a pathway between housing and health of ASR in high-income countries. By conducting a systematic review, we minimized the main problem of ad hoc search and selection of research findings, namely bias [59]. Adherence to the PRISMA checklist improved the reporting process and ensured a systematic and transparent approach [17]. The broad range of databases searched can also be seen as a strength. By not specifying a particular concept of housing, we avoided bias that might have resulted from the rejection of potentially relevant studies. To counteract selection bias, we did not restrict the search in terms of language or availability of the full text. In contrast to the review of Ziersch and Due [11], who drew on a wide range of refugee shelters, our study focused only on resettlement in HIC. By doing so, we reduced the contextual heterogeneity of the studies. Finally, a clear definition of psychosocial attributes according to Dunn [14] contributed to a systematic search within the studies, which in turn strengthens the reproducibility of our findings.

### Conclusions and recommendations for action

Previous research studies have shown that housing is an important social determinant of health. The literature provides evidence that psychosocial attributes address the social meanings of housing and create a sense of home, thus influencing the relationship between housing and health. Our study confirms this evidence on the influence of psychosocial attributes of housing on health for the population of ASR.

To conclude, housing in the context of health needs to be considered beyond its purely material/physical structures. Such structures are closely associated with psychosocial attributes which make homes meaningful, in particular the perceived degree of control over the home. The psychosocial attributes of housing derived by Dunn (2002) are of substantial relevance for the health of ASR, especially for their mental health. As psychosocial attributes are associated with the material/physical aspects of housing, they should never be considered alone but always together with the material/physical attributes. Finally, our findings confirm that the housing situation of ASR needs to be improved so that they can live healthier and more anxiety-free lives, become better integrated, and more satisfied.

On this basis, we recommend that housing for ASR should offer sufficient space and privacy, as well as opportunities to develop. Affordable housing is an important mental health issue and should hence be given high priority. Mass shelters, in which ASR often spend long periods of time, are associated with perceived insecurity and a lack of opportunities for retreat and development, which in turn affects their mental health and well-being. As a short-term response, opportunities for more privacy must be established until private accommodation becomes available, and social support services need to be strengthened. Furthermore, the area in which ASR are housed also plays an important role in their physical and mental health and well-being. ASR are often housed in disadvantaged areas with above-average crime rates, increasing the fear of being victimized. In such areas, neighborhood social support is often weak, so its role as an important mediator between housing and health that can promote ASR integration is lost. Accordingly, attention must be paid to neighborhood conditions when allocating housing to ensure healthy, anxiety-free living and successful integration.

Further research should examine the associations between psychosocial attributes and the health of ASR more closely. The interaction of the attributes with one another should be investigated. In addition, quantitative studies need to cover psychosocial attributes more comprehensively as qualitative findings demonstrate their relevance. However, due to the strong interconnectedness with the material/physical attributes of housing, psychosocial attributes should not be investigated detached from material/physical attributes.

## Data Availability

All data produced in the present work are contained in the manuscript.

## Abbreviations

ASR: Asylum seekers and refugees

## Authors’ contributions

TB is the guarantor of the review. OR and OS developed the study idea and obtained funding. VD importantly contributed to the idea, design, and conceptualization. VD and TB conducted screening, data extraction, and quality assessment. TB drafted the manuscript; VD and OR significantly contributed to the writing. OS and OR revised various versions of the manuscript critically for intellectual content. All authors read and approved the final version of the manuscript.

## Availability of data and materials

Not applicable.

## Ethics approval and consent to participate

Not applicable. As this review does not collect primary data from individuals but performs a systematic literature search, it did not need to be approved by an ethics committee.

## Consent for publication

Not applicable.

