## Supplementary material for "Psychosocial attributes of housing and their relationship with health among refugee and asylum-seeking populations in high-income countries: systematic review": Supplemenary Files

### Supplementary file 1: Indicators of psychosocial attributes derived from Dunn (2002; own representation) (Psychosocial attributes of housing and health among refugees, systematic review, high-income countries, 1995-2022)

| | | |
|---|---|---|
| 1 | **Measures of demand** | |
| 1.1 | Housework strain | Degree to which participants perceive any work associated with the home (including gardening) physically or mentally as a strain or burden |
| 1.2 | Strain of meeting costs | Degree to which participants perceive housing costs (including affordability issues) physically or mentally as a strain or burden. (Note: not affordability itself but the individual burden of affordability issues is addressed here) |
| 2 | **Measures of control** | |
| 2.1 | Place of refuge | Degree to which participants feel uncomfortable at home (such as issues of disturbed privacy, lack of retreat or low opportunities to personalize own home) |
| 2.2 | Worry of forced move | Degree to which participants are worried about a forced move (demonstrating a lack of control of stable housing). Note: not the number of forced moves but the feelings about it (such as worry, fear) are addressed here |
| 2.3 | Worry of frequent moves | Degree to which participants perceive high housing mobility (frequent moves) as a burden or strain. Note: not the frequency of moves itself but the perceptions about it are addressed here |
| 2.4 | Fear of crime / victimization | Degree to which participants feel unsecure at home and fear becoming a victim of crime: security / safety issues in the domestic environment (home including neighbourhood) |
| 3 | **Measures of expressing status** | |
| 3.1 | Pride | Degree to which participants are proud to show their homes to visitors |
| 3.2 | Self-reflection | Degree to which participants feel like their homes reflects who they are |
| 3.3 | Belonging | Degree to which participants feel like they belong in their neighbourhood |
| 4 | **General measures** | |
| 4 | Satisfaction | Degree to which participants are satisfied with their housing situation (different components of housing are possible here, as long as it relates to housing / home) |

### Supplementary file 2: Search terms (Psychosocial attributes of housing and health among refugees, systematic review, high-income countries, 1995-2022)

**PubMed**

((((((("housing"[Title/Abstract] OR "accommodation"[Title/Abstract] OR "dwelling"[Title/Abstract] OR "shelter"[Title/Abstract] OR "home"[Title/Abstract]) NOT ("ocular"[All Fields] OR "oculars"[All Fields])) NOT ("animals"[MeSH Terms:noexp] OR "animal"[All Fields])) NOT "homeless"[All Fields]) NOT ("homeless persons"[MeSH Terms] OR ("homeless"[All Fields] AND "persons"[All Fields]) OR "homeless persons"[All Fields] OR "homeless"[All Fields] OR "homelessness"[All Fields])) OR ("housing/adverse effects"[MeSH Terms] OR "housing/classification"[MeSH Terms] OR "housing/instrumentation"[MeSH Terms] OR "housing/methods"[MeSH Terms] OR "housing/standards"[MeSH Terms] OR "housing/trends"[MeSH Terms])) NOT "housing, animal"[MeSH Terms]) AND ("humans"[MeSH Terms] AND 1995/01/01:2022/04/20[Date - Publication])

AND

(("health"[Title/Abstract] OR "morbidity"[Title/Abstract] OR "mortality"[Title/Abstract] OR "depression"[Title/Abstract] OR "post-traumatic stress"[Title/Abstract] OR "posttraumatic stress"[Title/Abstract] OR "anxiety"[Title/Abstract]) AND "humans"[MeSH Terms] AND ("humans"[MeSH Terms] AND 1995/01/01: 2022/04/20[Date - Publication]))

AND

(("refugee*"[All Fields] OR ("asylum"[All Fields] OR "asylum s"[All Fields] OR "asylums"[All Fields]) OR "forced migration"[All Fields] OR "forced migrant*"[All Fields] OR "displaced person*"[All Fields] OR "displaced population*"[All Fields] OR "refugees"[MeSH Terms]) AND ("humans"[MeSH Terms] AND 1995/01/01: 2022/04/20[Date - Publication]) AND "humans"[MeSH Terms]) AND "humans"[MeSH Terms]) AND (humans[Filter])

**Web of Science**

(TI=hous* OR TI=accommodation* OR TI=dwelling* OR TI=shelter* OR AB=hous* OR AB=accommodation* OR AB=dwelling* OR AB=shelter* NOT ALL=ocular NOT ALL=oculars NOT ALL=homeless NOT ALL=homelessness)

AND

(TI=health OR TI=morbidity OR TI=mortality OR TI=depression OR TI=anxiety OR TI="post traumatic stress" OR AB=health OR AB=morbidity OR AB=mortality OR AB=depression OR AB=anxiety OR AB="post traumatic stress")

AND

(ALL=refugee* OR ALL=asylum OR ALL=("forced migration") OR ALL=("forced migrant*") OR ALL=("displaced person*") OR ALL=("displaced population*"))

**CINAHL, SOCIndex, PSYCHIndex, PSYCHINfo**

"((TI (hous* OR dwelling* OR shelter* OR accommodation*)) OR (AB (hous* OR dwelling* OR shelter* OR accommodation)) NOT (ocular OR homeless OR homelessness))

AND

((TI (health OR morbidity OR mortality OR depression OR "post-traumatic stress" OR "posttraumatic stress" OR anxiety)) OR (AB (health OR morbidity OR mortality OR depression OR "post-traumatic stress" OR "posttraumatic stress" OR anxiety)))

AND

((TX refugee*) OR (TX displaced person*) OR (TX displaced population*) OR (TX forced migration) OR (TX forced migrant*) OR (TX asylum seeker*) OR (TX asylum-seeker*)) Published Date: 19950101-20210420"

**Cochrane Library**

(housing OR dwelling OR accommodation OR shelter):ti,ab,kw NOT (homeless OR homelessness OR ocular) OR MeSH descriptor: [Housing] explode all trees NOT MeSH descriptor: [Housing, Animal] explode all trees

AND

(health):ti,ab,kw OR (morbidity OR mortality):ti,ab,kw OR ("post-traumatic stress"):ti,ab,kw OR (depression):ti,ab,kw OR (anxiety):ti,ab,kw

AND

(refugee):ti,ab,kw OR ("asylum seeker"):ti,ab,kw OR ("forced migration" OR "forced migrant"):ti,ab,kw OR ("displaced person" OR "displaced population"):ti,ab,kw OR MeSH descriptor: [Refugees] explode all trees

**Supplementary file 3: Conceptual map (Psychosocial attributes of housing and health among refugees, systematic review, high-income countries, 1995-2022)**

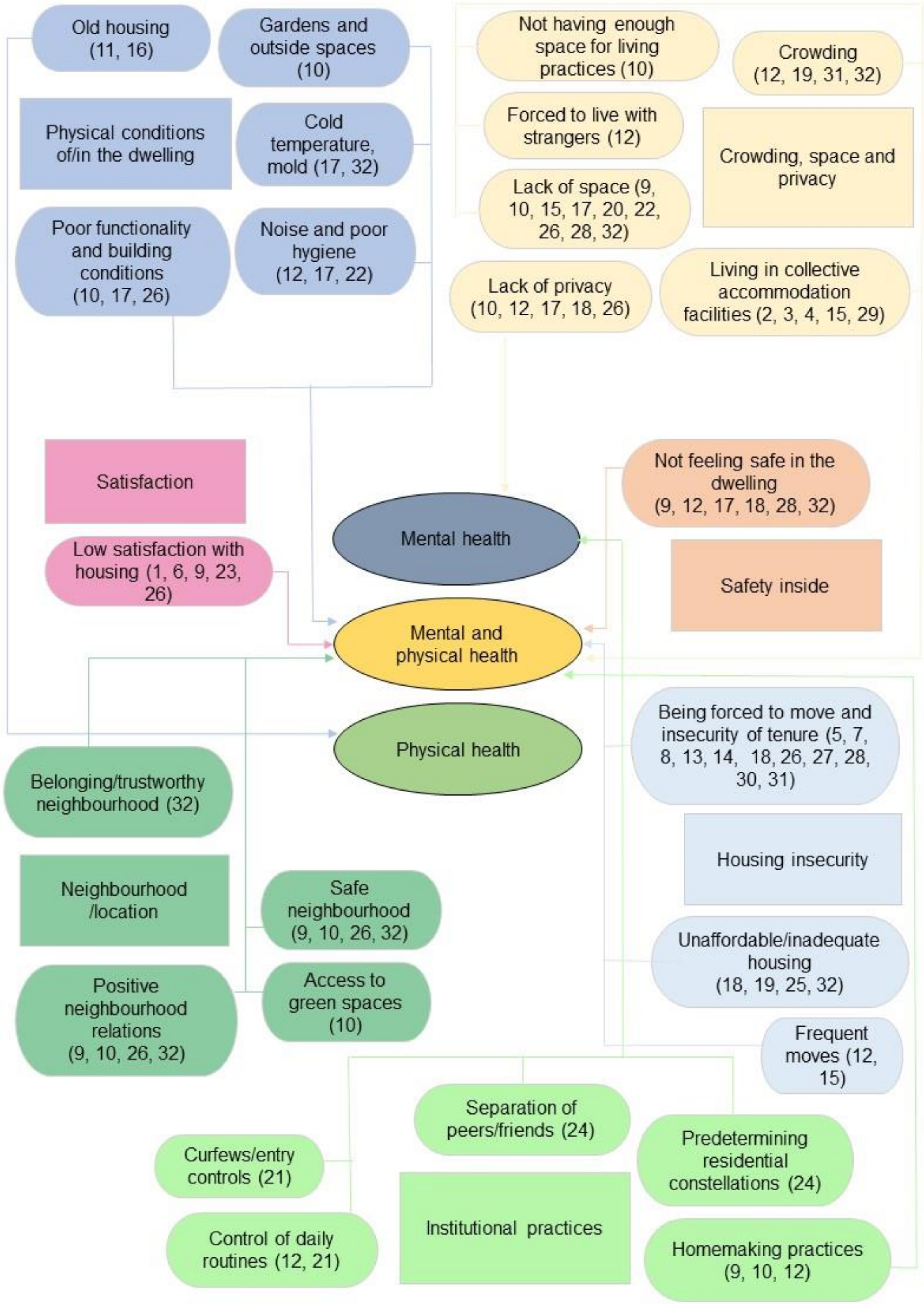

### Supplementary file 4: Characteristics of studies included (Psychosocial attributes of housing and health among refugees, systematic review, high-income countries, 1995-2022)

| No. | Reference | Study design | Study population | Study region<br>1: resettlement country<br>2: origin of participants | Exposure<br>1: assessed housing components<br>2: assessed psychosocial attributes | Outcome | MMAT Score |
|---|---|---|---|---|---|---|---|
| 1 | Ahmad et al. (2020) | Quantitative: findings from a Canadian longitudinal study: Syrian Refugee Integration and Long-term Health Outcomes in Canada study (SyRIA.lth) | 1924 refugees recruited through a variety of community-based strategies, arriving between January 2015 and June 2017 | 1: Canada<br>2: Syria | 1: satisfaction with housing<br>2: general measures | Severe- and/or moderate-level of depression symptoms at year-2 (Nine-item Patient Health Questionnaire (PHQ-9) with a score range of 0-27) | **** |
| 2 | Ambrosetti et al. (2021) | Quantitative: linear regression and panel models with longitudinal data from IAB-BAMF-SOEP survey | 3975 adult ASR who arrived in Germany for humanitarian reasons between 2013 and 2016 and registered in the AZR (Ausländerzentralregister) by January 2017 | 1: Germany<br>2: Syria, Afghanistan, Eritrea, Iraq | 1: type of residence<br>2: N.A. | Life satisfaction and self-rated health | **** |
| 3 | Bakker et al. (2016) | Quantitative; Comparison of NL and UK; NL: Survey integration for new groups (SING09): cross-sectional dataset, UK: Survey of New Refugees (SNR): longitudinal study of refugee integration: 4 waves: 1 one week after leave to remain granted, 2 after 8, 3 after 15 and 4 after 21 | Refugees; NL: focus on four largest refugee groups, total: 2980 refugees, UK: all new refugees over 18 who were granted leave (temporary or indefinite), last wave: 921 refugees | 1: Netherlands and UK<br>2: Afghanistan, Iraq, Iran, Somalia | 1: Type of accommodation during the asylum procedure<br>2: measures of control | Poor physical and mental health: operationalization of both: (1) Very bad to (5) very good | *** |

| No. | Reference | Study design | Study population | Study region<br>**1: resettlement country**<br>**2: origin of participants** | Exposure<br>**1: assessed housing components**<br>**2: assessed psychosocial attributes** | Outcome | MMAT Score |
|---|---|---|---|---|---|---|---|
| 4 | Bakker et al. (2014) | Quantitative: Large-scale dataset: SING2009 was used: cross-sectional | 2907 refugees with flight experience and motive, who stay/ed in asylum acc., age 16 to 65 | 1: Netherlands<br>2: Afghanistan, Iraq, Iran, Somalia | 1: length of stay in state-provided asylum accommodation/centres<br>2: N.A. | Poor mental health (SF-12) | **** |
| 5 | Bhui et al. (2012) | Quantitative: cross-sectional | 142 refugees between 18 and 65; 50 recruited from GP's registers and remaining 100 from non-conventional places (e.g. cafes, community centres, etc.); recruited when Somali origin, gave informed consent and were resident in the London Boroughs or Lambeth at time of survey | 1: United Kingdom<br>2: Somalia | 1: residential mobility<br>2: N.A. | Psychiatric disorder (Mini Neuropsychiatric Interview (MINI)). General health (first question of Short Form-12) | **** |
| 6 | Campbell et al. (2018) | Quantitative: Prospective longitudinal cohort study | 5678 new adult refugees, identified by the Border Agency's central database, from 1st Dec.-25th March 2007, one week after they were granted refugee status and leave to remain in the UK: Longitudinal Survey of New Refugees (4 waves: 1 one week after leave to remain granted, 2 after 8, 3 after 15 and 4 after 21 months) | 1: United Kingdom<br>2: Afghanistan, Africa, Iran, Pakistan, Iraq, USA, Turkey, Europe | 1: satisfaction with current accommodation (just measured in 1 and 2 wave)<br>2: general measures | Poor mental health and emotional well-being Emotional wellbeing (self-reported 5-point ordinal scale of SF-36 Health Survey Questionnaire) | **** |

| No. | Reference | Study design | Study population | Study region<br>1: resettlement country<br>2: origin of participants | Exposure<br>1: assessed housing components<br>2: assessed psychosocial attributes | Outcome | MMAT Score |
|---|---|---|---|---|---|---|---|
| 7 | Cooperi et al. (2019) | Quantitative: cohort study | 2399 refugees | 1: Australia<br>2: Middle East, Central Asia, Southern Asia, Africa, South-East Asia | 1: type of housing contract<br>2: N.A. | Two dichotomous outcome measures: High Risk of Severe Mental Illness (HR-SMI) via Kessler-6; PTSD via PTSD-8 | **** |
| 8 | Correa-Velez et al. (2012) | Quantitative: longitudinal analysis | 120 resettled refugee youth typically spend 6-12 months at an English Language School in Australia before entering mainstream schools | 1: Australia<br>2: Africa, Middle East, Eastern Europe, Southeast Asia | 1: moving house over the previous year<br>2: N.A. | Subjective health status ("How satisfied are you with your health?", 5-point scale based on WHO, 1996) | *** |
| 9 | Dudek et al. (2022) | Quantitative: explorative cluster analysis of population-based, cross-sectional secondary data to identify clusters of refugee accommodation and mixed model analysis; data: IAB-BAMF-SOEP survey | 1535 adult ASR | 1: Germany<br>2: Afghanistan, Syria, Iraq, Eritrea, stateless, other countries | 1: area type, security in neighbourhood, accessibility of public transport, leisure time activities; dwelling type, number of residents per unit/household, number of rooms, furnishment, inviting friends, time spent in boredom (hours per week), satisfaction with living situation, satisfaction with allocated place of residence, satisfaction with size of the dwelling, sense of belonging, free choice of residence, receiving housing allowance, paying rent<br>2: measures of control, expressing status, general measures | Mental health and physical health (SF-12) | ***** |

| No. | Reference | Study design | Study population | Study region<br>**1: resettlement country**<br>**2: origin of participants** | Exposure<br>**1: assessed housing components**<br>**2: assessed psychosocial attributes** | Outcome | MMAT Score |
|---|---|---|---|---|---|---|---|
| 10 | Due et al. (2020) | Qualitative: photovoice study | 11 adult ASR | 1: Australia<br>2: Africa, Middle East, South-East Asia | 1: range of different housing concerns emerged from interviews: gardens and outside spaces, housing conditions, layout/space/privacy, furnishing and home-making practices, neighbourhoods' location, safety<br>2: measures of demand, control, expressing status | Self-reported health and wellbeing, level of ontological security | ***** |
| 11 | Eisenberg et al. (2011) | Quantitative: longitudinal analysis | 1148 refugee children younger than 7 years who arrived in Massachusetts from 2000 to 2007 | 1: United Stated<br>2: Europe, Central Asia, Africa, East Asia, and Pacific, Near East and South Asia, Latin America, and the Caribbean | 1: age of housing<br>2: N.A. | High blood lead levels | ***** |
| 12 | Gewalt et al. (2019) | Qualitative: interviews | 21 women seeking asylum in pregnancy and early motherhood | 1: Germany<br>2: West Africa, East Europe, Asia | 1: stressful living conditions: high background noise, crowding / lack of privacy (sharing rooms with strangers), control of daily routines, safety issues (fear of verbal and physical violence in acc.), frequent transfers<br>2: measures of control | Perceived health and wellbeing | ***** |

| No. | Reference | Study design | Study population | Study region<br>**1: resettlement country**<br>**2: origin of participants** | Exposure<br>**1: assessed housing components**<br>**2: assessed psychosocial attributes** | Outcome | MMAT Score |
|---|---|---|---|---|---|---|---|
| 13 | Gillespie et al. (2020) | Quantitative: Somali Youth Longitudinal Study (SYLS); data at Time 3 and Time 4 was analysed | Refugees; had to have lived in the U.S. or Canada for at least one year, to be either from Somalia or of Somali descent (i.e., children of Somali refugees born outside of Somalia), and be between the ages of 18 and 30; 265 participants at time 3, 198 at time 4 | 1: United States<br>2: Somalia | 1: residential mobility<br>2: measures of demand, control, expressing status | Post-traumatic stress (Harvard Trauma Questionnaire, HTQ, + mean score to summarize variable) | ***** |
| 14 | Hocking (2018) | Report on qualitative data derived from a prospective, mixed methods study design | 56 current and former community-dwelling ASR | 1: Australia<br>2: Sri Lanka, Pakistan, Zimbabwe, Iraq, Afghanistan | 1: stable housing<br>2: measures of control | Self-perceived emotional health | ***** |
| 15 | Hynek et al. (2018) | Qualitative: interviews | 6 Scientists and 5 Practitioners about UMR | 1: Austria<br>2: N.A. | 1: housing situation: living arrangements and conditions of UMRs, including type of accommodation (living with a foster family or in a residential group, a residential home, supervised accommodation), and the number of roommates and reallocations<br>2: measures of control | Mental health (risk factors) | **** |

| No. | Reference | Study design | Study population | Study region<br>**1: resettlement country**<br>**2: origin of participants** | Exposure<br>**1: assessed housing components**<br>**2: assessed psychosocial attributes** | Outcome | MMAT Score |
|---|---|---|---|---|---|---|---|
| 16 | Kotey et al. (2018) | Quantitative: cross-sectional study | 1950 refugee children younger than 15 years | 1: United States<br>2: Asia, Eurasia, Latin America, and Caribbean, Middle West and Sub-Saharan Africa | 1: age of housing<br>2: N.A. | Elevated Blood Lead Levels and intestinal infestation | *** |
| 17 | Lephard & Haith-Cooper (2016) | Qualitative: semi-structured interviews | 6 Asylum seeking women: 4 in the UK longer than a year before becoming pregnant, 2 arrived partway through their pregnancy | 1: United Kingdom<br>2: Sub-Saharan Africa, Eastern Europe | 1: Inappropriate accommodation in terms of: space, hygiene, temperature, physical barriers, insecurity, privacy (forced to live with strangers), smell<br>2: measures of control | Perceived physical and mental health | **** |
| 18 | Mangrio et al. (2020) | Qualitative descriptive study (thematic network analysis) | 24 newly arrived refugee parents | 1: United States<br>2: Syria | 1: Trying to find stable housing and associated issues of privacy, safety, residential mobility, housing costs<br>2: measures of demand, control | | **** |
| 19 | Miller et al. (2002) | Qualitative: semi-structured interviews | 28 adult Bosnian refugees participating in a mental health program | 1: United States<br>2: Bosnia | 1: Crowding (big families with little space) and affordability<br>2: measures of demand, control | Exile-related distress and recovery from war-trauma | ***** |

| No. | Reference | Study design | Study population | Study region<br>**1: resettlement country**<br>**2: origin of participants** | Exposure<br>**1: assessed housing components**<br>**2: assessed psychosocial attributes** | Outcome | MMAT Score |
|---|---|---|---|---|---|---|---|
| 20 | Müller et al. (2020) | Quantitative: retrospective cross-sectional study | 1957 minor ASR residing in two large reception centres in Northern Germany | 1: Germany<br>2: Syria, Iraq, Afghanistan, Georgia, other | 1: residential density<br>2: N.A. | Acute respiratory infections according to ICD-10 codings of acute respiratory infections | ***** |
| 21 | Murphy et al. (2018) | Qualitative: narrative study | 16 adult asylum seekers with experiences of attending mental health services | 1: Ireland<br>2: Africa | 1: "institutional practices": curfews and entry controls, daily decisions decided by management<br>2: measures of control, expressing status | Perceived meaning in life as part of a positive mental health state. | **** |
| 22 | Nikolai et al. (2020) | Qualitative: free-listing, open-ended interviews | 30 adult ASR living in Switzerland since 2011 | 1: Switzerland<br>2: Syria | 1: housing problems (structural and socio-cultural problems): adverse/crowded conditions (with consequences regarding noise, hygiene, lack of space, family conflicts)<br>2: measures of control | Self-reported psychological or emotional problems and self-reported physical health problems | ***** |
| 23 | Nutsch & Bozorgmehr (2020) | Quantitative: secondary data regression analysis, cross-sectional | 4465 ASR | 1: Germany<br>2: Syria, Iraq, Afghanistan | 1: housing satisfaction<br>2: general measures | Depression (Patient Health Questionnaire 2, PHQ-2) | ***** |
| 24 | Omland & Andenas (2020) | Qualitative: semi-structured interviews at two time points in two accommodation types for UAM | 15 UMR aged 13 to 16 (+ their caregivers) living in Norway for 3-5 months and residing in care centres at time of first interview | 1: Norway<br>2: Afghanistan, Somalia, Angola, Sri Lanka | 1: Relational practices in accommodation facilities (such as separating friends, avoiding similar cultural groups or the development of a collective conduct)<br>2: measures of control, expressing status | Facilitating a sense of togetherness to promote the health of UMR | *** |

| No. | Reference | Study design | Study population | Study region<br>**1: resettlement country**<br>**2: origin of participants** | Exposure<br>**1: assessed housing components**<br>**2: assessed psychosocial attributes** | Outcome | MMAT Score |
|---|---|---|---|---|---|---|---|
| 25 | Palmer & Ward (2007) | Qualitative: semi-structured interviews | 21 ASR (6 AS, 1 on exceptional leave, 1 with permanent residence, 13 refugees) | 1: United Kingdom<br>2: Africa, Middle East, Eastern Europe, South America | 1: lack adequate housing<br>2: N.A. | Perceived mental health | **** |
| 26 | Papadopoulous et al. (2004) | Qualitative: semi-structured interviews | 106 ASR (47% permanent residency or refugee status, 36% temporally admitted, 12% on exceptional leave) | 1: United Kingdom<br>2: Ethiopia | 1: Satisfaction with accommodation, housing problems (space, privacy), conditions of the building, neighbourhood relations, security of tenancy, safety<br>2: measures of expressing status, general measures | Meanings of health | ***** |
| 27 | Sundell et al. (2016) | Quantitative: cross-sectional analysis of survey data | 587 adult refugees with residence permit for 2-5 months | 1: Sweden<br>2: Iraq | 1: housing problems (housing stability)<br>2: N.A. | Mental health (dichotomous variable of the 12-item version of General health Questionnaire, GHQ-12) | **** |
| 28 | Vitale & Rhyde (2016) | Qualitative: semi-structured interviews | 9 adult male refugees having been granted refugee status within two years | 1: United Kingdom<br>2: Middle East, Africa | 1: Access to adequate accommodation, safety, space, mobility<br>2: measures of control | Self-reported mental health and wellbeing | ***** |
| 29 | Walther et al. (2020) | Quantitative: cross-sectional secondary data regression analysis | 4325 adult ASR who arrived in Germany between 2013 and 2016 | 1: Germany<br>2: Syrian, Afghan, Iraqi, Eritrean, other | 1: living in collective facilities<br>2: N.A. | Psychological distress (Patient Health Questionnaire for Depression and Anxiety, PHQ-4) | ***** |

| No. | Reference | Study design | Study population | Study region<br>**1: resettlement country**<br>**2: origin of participants** | Exposure<br>**1: assessed housing components**<br>**2: assessed psychosocial attributes** | Outcome | MMAT Score |
|---|---|---|---|---|---|---|---|
| 30 | Warfa et al. (2006) | Qualitative: discussion groups | 13 Somali professionals (two discussion groups) and 21 Somali lay people (in four groups) | 1: United Kingdom<br>2: Somalia | 1: residential mobility<br>2: measures of control | Core themes of mental health emerged in the interviews: stress, distress, worry and anxiety | ***** |
| 31 | Whitsett et al. (2017) | Quantitative: longitudinal analysis of survey data | 105 treatment seeking adult asylum seekers being survivors of torture | 1: United States<br>2: Africa, Middle East, South Asia | 1: housing status<br>2: N.A. | PTSD (part IV of the HTQ), general psychiatric symptoms (HSCL) | *** |
| 32 | Ziersch et al. (2017) | Qualitative: semi-structured interviews | 50 ASR purposely sampled from broader sample | 1: Australia<br>2: Africa, Middle East, South-East Asia | 1: affordability, physical elements, (cold, damp, space, layout), social elements (feeling of safety and belonging, interaction with neighbours), insecurity of tenure<br>2: measures of demand, control, expressing status | Mental health: self-described symptoms of anxiety, depression, and stress, (minor topic:) suicidal ideation. Physical health: chronic pain, sleep disturbances | **** |

### Supplementary file 5: Thematic analysis (Psychosocial attributes of housing and health among refugees, systematic review, high-income countries, 1995-2022)

| Housing themes |
|---|
| Crowding, space, privacy (including collective facilities) |
| Security of tenancy, affordability and residential mobility |
| Safety |
| Physical conditions of / in the dwelling |
| Satisfaction |
| Social relations (neighborhood, family, other residents) |
| Practices inside accommodation |

| Psycho-social attributes |
|---|
| described by study participants |
| quantitatively assessed |
| derived indirectly from study results |

| Health topics |
|---|
| Mental health |
| Physical health |
| Both |

| Authors | Housing components relevant to health | Health Outcome | Psycho-social attributes |
|---|---|---|---|
| Ahmad et al. (2020) | Housing satisfaction | Severe- and/or moderate-level of depression symptoms at year-2 | 4. Satisfaction |
| Ambrosetti et al. (2021) | Living in shared accomodation | Life satisfaction and self-reported health | N.A. |
| Bakker, Cheung & Phillimore (2016) | Living in state-provided asylum accomodation | Poor physical and mental health | 2.1 Place of refuge |
| Bakker, Dagevos & Engbersen (2014) | Stay in asylum accomodation for 5 years or longer | Poor mental health | N.A. |
| Bhui et al. (2012) | Residential mobility | Psychiatric disorder | N.A. |
| Campbell et al. (2018) | Satisfaction with current accomodation | Poor mental health and emotional well-being | 4. Satisfaction |
| Cooperi et al. (2019) | Type of housing contract | Severe mental illness | N.A. |
| Correa-Velez, Gifford & McMichael (2015) | Residential mobility | Subjective health status and well-being | N.A. |
| Dudek, V., Razum, O. & Sauzet, O. (2022) | Type of dwelling and area, number of rooms and residents, living space (sqm)<br>Safety (in acc.)<br>Satisfaction with current accomodation, place of residence and living situation<br>Furnishing and home-making practices<br>Neighborhood relations | Assessed physical and mental health | 2.1 Place of refuge<br>2.4 Fear of crime/victimization<br>3.3 Belonging<br>4. Satisfaction |
| Due et al. (2020) | Gardens and outside spaces<br>Housing conditions<br>Layout, space, privacy<br>Furnishing and home-making practices<br>Neighborhood location and relations<br>Safety | Self-reported health and well-being | 1.1 Housework strain<br>1.2 Strain of meetings costs<br>2.1 Place of refuge<br>2.2 Worry of forced move<br>2.4 Fear of crime/victimization<br>3.3 Belonging |

| | | | | |
|---|---|---|---|---|
| Eisenberg et al. (2011) | Age of housing | | High blood lead levels | N.A. |
| Gewalt et al. (2019) | Constant backgorund noise | | Perceived health and wellbeing | 2.1 Place of refuge |
| | Sharing rooms with strangers | | | 2.4 Fear of crime/victimization |
| | Control of daily routines/type of food | | | |
| | Fear of verbal and physical crime | | | |
| | Frequent transfers | | | |
| Gillespie et al. (2020) | Residential mobility/Housing stability | | Post-traumatic stress | 2.4 fear of crime |
| | | | | 3.3 Belonging |
| Hocking (2018) | Residential mobility (stable housing) | | Self-perceived emotional health | 2.3 Worry of frequent move |
| Hynek et al. (2020) | Living conditions | | Poor mental health | 2.3 Worry of frequent moves |
| | Type of accommodation | | | |
| | Number of roommates | | | |
| | Number of reallocations | | | |
| Kotey et al. (2018) | Age of housing | | Elevated Blood Lead Levels and intestinal infestation | N.A. |
| Lephard & Haith-Cooper (2016) | Space | | Perceived physical and mental health | 2.1 Place of refuge |
| | Hygiene | | | 2.4 fear of crime |
| | Temperature (cold rooms) | | | |
| | Physical barriers | | | |
| | Safety (in acc.) | | | |
| | Privacy | | | |
| | Smell | | | |
| Mangrio et al. (2020) | Privacy | | Stress causing mental struggle | 1.2 strain of meeting costs |
| | Safety (in acc.) | | | 2.1 place of refuge |
| | Residential mobility | | | 2.3 frequent moves |
| | Housing costs | | | 2.4 fear of crime |
| Miller et al. (2002) | Crowding | | Stressors impeding recovery from war-trauma | 1.2 strain of meeting costs |
| | Affordability | | | 2.1 place of refuge |
| Müller et al. (2020) | Residential density | | Risk of respiratory infections | N.A. |
| Murphy et al. (2018) | Institutional practices | Curfews | Perceived meaning of life (mental health) | 2.1 place of refuge |
| | | exit / entry controls | | 3.1 pride |
| | | Agency over daily decisions | | 3.2 self-reflection |
| Nikolai et al. (2020) | Noise | | Psychological stress (space) | 2.1 place of refuge |
| | Hygiene | | Physical health problems (hygiene) | |
| | Lack of space | | | |
| | Family conflicts | | | |
| Nutsch & Bozorgmehr (2020) | Housing satisfaction | | Depression | 4. satisfaction |
| Omland & Andenas (2020) | Relational practices | separating friends | Sense of togetherness / developing strategies to promote mental health | 2.1 place of refuge |
| | | avoiding similar cultural groups | | 3.3 sense of belonging |
| | | developing collective conduct | | |
| Palmer & Ward (2007) | Lacking adequate housing | | Stress and perceived mental health | N.A. |
| Papadopoulous et al. (2004) | Satisfaction | | Meanings of health (physical and mental well-being) | 3.3 Sense of belonging |
| | Space | | | 4. Satisfaction |
| | Privacy | | | |
| | Neighborhood relations | | | |
| | Building conditions | | | |
| | Security of tenancy | | | |
| | Safety in neighbourhood | | | |
| Sundell et al. (2016) | Housing stability | | Mental health (GHQ-12) | N.A. |
| Vitale & Rhyde (2016) | Housing access | | Self-reported mental health and well-being | 2.1 place of refuge |
| | Safety | | | 2.2 forced moves |
| | Space | | | |
| | Mobility | | | |

| | | | |
|---|---|---|---|
| Walther et al. (2020) | Living in collective facilities | Psychological distress (PHQ-4) | N.A. |
| Warfa et al. (2006) | Residential mobility | Mental health (core themes: distress, worry, anxiety) | 2.1 place of refuge<br>2.2 forced moves<br>2.3 frequent moves |
| Whitsett et al. (2017) | Instable accommodation<br>Crowded accommodation | PTSD (HTQ), psychiatric symptoms (HSCL) | N.A. |
| Ziersch et al. (2017) | Affordability<br>Temperature and moisture<br>Space<br>Overcrowding<br>Safety (in acc.)<br>Belonging<br>Social relations<br>Insecurity of tenure | Self-described symptoms of anxiety, depression and stress; self-described physical health problems (chronic pain, sleep disturbances) | 1.2 strain of meeting costs<br>2.1 place of refuge<br>2.2 worry of forced move<br>2.4 fear of crime<br>3.3 sense of belonging |
